# Recovery in pupillometric non-visual functions following chiasmal decompression in pituitary adenoma

**DOI:** 10.1101/2025.10.22.25338480

**Authors:** Daniella Mahfoud, Jensen Ang, Beng Ti Ang Christopher, Monisha Esther Nongpiur, Dan Milea, Raymond P. Najjar

**Author notes:** Corresponding Author: Dr Raymond P. Najjar Department of Ophthalmology, Yong Loo Lin School of Medicine, National University of Singapore, Singapore 119228, Singapore. These authors contributed equally to this work (Daniella Mahfoud, Jensen Ang).

## Abstract

**Background:** This study investigates visual and non-visual (*i.e.,* pupillometric) changes associated with optic nerve compression and decompression, and the potential role of chromatic pupillometry as an objective evaluation tool for visual dysfunction in pituitary adenoma (PA).

**Methods:** This longitudinal study included 27 patients with PA (median age = 55.4 years [IQR=23.4]) and 41 age-matched controls (53.6 [13.7] years), evaluated pre-operatively (7.0 ± 5.4 weeks prior to surgery) and post-operatively (12.0 ± 1.5 weeks post-surgery) with handheld chromatic pupillometry, in addition to comprehensive neuro-ophthalmological and neuroimaging examinations. Pupillometric features were analyzed for associations with structural changes and visual outcomes.

**Results:** Transsphenoidal surgery significantly reduced the upward displacement and increased the thickness of the optic chiasm measured on MRI (p<0.001). Pupillary responses improved post-operatively, including increased maximum constriction to both red and blue light, but remained below control levels (p<0.05). In patients with full visual field index (VFI) recovery (n=5), pupillometric responses were comparable to controls (p>0.05) while patients without recovery exhibited persistent deficits in these metrics. Remarkably, melanopsin-driven post-illumination pupil responses (PIPR) improved significantly, reaching control values post-operatively (p>0.05), regardless of VFI recovery. Structural recovery correlated with improved VFI (ρ= -0.62, p<0.001) and with maximum constriction to red light (ρ= -0.47, p=0.004).

**Conclusions:** PA surgery leads to a consistent recovery in pupillary light responses that correlate with clinical structural and functional changes observed in PA patients. Notably, melanopsin-mediated responses normalized post-operatively even in patients without VFI recovery, suggesting that non-visual pathways can recover independently of vision.

**What is already known on this topic:** Pituitary adenomas frequently cause optic chiasm compression, leading to visual field loss and sleep disturbances. While transsphenoidal surgery can relieve the structural compression, functional recovery of visual and non-visual pathways remains variable and difficult to predict with existing clinical tests such as perimetry, OCT, or MRI.

**What this study adds:** In this prospective study, handheld chromatic pupillometry objectively detected recovery in pupillary light responses after pituitary adenoma surgery, in parallel with structural and visual improvements. Notably, melanopsin-driven post-illumination pupil responses normalized even in patients without visual field recovery, suggesting that non-visual pathways may recover independently of vision.

**How this study might affect research, practice or policy:** These findings establish handheld chromatic pupillometry as a sensitive, non-invasive biomarker to monitor both visual and non-visual functional outcomes after chiasmal decompression. Incorporating this approach into clinical and research protocols could improve postoperative monitoring and guide future work linking visual and systemic recovery in pituitary adenoma.

## INTRODUCTION

Pituitary adenomas (PAs) are common intracranial tumors that can compress the visual pathways, causing visual impairment in 30-70% of cases(1) and significantly affecting quality of life. Although transsphenoidal surgery (TSS) is the standard treatment for PAs(2), visual recovery following surgery varies widely and is not always predicted by anatomical restoration seen on magnetic resonance imaging (MRI)(3,4). Conventional evaluation methods such as automated perimetry, optical coherence tomography (OCT), diffusion-weighted MRI, and visual evoked potentials (VEP)(5,6), are often complex, time-consuming, or patient-dependent. While OCT has demonstrated strong diagnostic utility in detecting compressive chiasmopathy and predicting visual recovery based on retinal nerve fiber layer (RNFL) thinning(1,7), it primarily reflects structural damage rather than real-time neuro-ophthalmic function. Similarly, MRI delineates chiasmal displacement but lacks predictive power for functional outcomes(6). Perimetry relies on patient cooperation(8), and VEP, though objective, is expensive, technically demanding, and time-intensive, limiting its widespread clinical utility(9). These limitations highlight the need for a rapid, objective, and accessible method for tracking neuro-ophthalmic function in PA patients.

Quantitative pupillometry has emerged as a promising tool for assessing neuro-ophthalmic function(10,11) with studies showing improvement in indices like the neurological pupil index (NPi) following TSS, paralleling visual acuity gains(12,13). However, its potential for tracking visual field recovery and chiasmal plasticity remains unexplored. Chromatic pupillometry, which selectively stimulates different photoreceptor pathways, offers added functional insights. Rods and cones mediate transient pupillary constriction, while intrinsically photosensitive retinal ganglion cells (ipRGCs) drive sustained responses, particularly to blue light(14,15). These distinct response patterns allow for a functional assessment of retinal and optic nerve integrity, which has been demonstrated in patients with neuro-ophthalmic conditions(15–18). Importantly, because ipRGC-driven responses also underpin non-visual functions such as circadian regulation and sleep(19), chromatic pupillometry offers insights into physiological functions that extend beyond vision (i.e., sleep quality), which may also be disrupted in patients with PA(20). For instance, the post-illumination pupil response (PIPR), largely driven by melanopsin-expressing ipRGCs, is a robust marker of non-visual light responses(21,22) and may reveal differential recovery trajectories that are not apparent through conventional ophthalmic tests. Investigating these pathways could expand our understanding of PA surgery outcomes and provide a functional biomarker of visual recovery as well as systemic health implications.

In this study, we used handheld chromatic pupillometry (HCP) to investigate dynamic pupillometric features before and after TSS and their relationships with visual outcomes. We hypothesize that improvements in pupillary responses, such as maximum constriction amplitudes and PIPRs, will parallel structural and functional recovery, providing an objective indicator of both neuro-ophthalmic and non-visual outcomes following surgical decompression.

## METHODS

### Participants

This prospective, longitudinal study included a total of 27 patients with PA and 41 healthy controls. Patients were recruited from the Neurosurgery and Neuro-Ophthalmology clinics at Singapore General Hospital (SGH), while healthy controls were recruited from SGH General Ophthalmology clinics or the general population. Variations in sample size, reported throughout the manuscript, were due to some patients having performed some of the assessments at other hospitals or missing data (Supplementary Figure 1). Eligibility criteria are reported in supplementary material.

### Study assessments

All patients underwent pre-operative (7.0±5.4 weeks before surgery) and post-operative (12.0±1.5 weeks after surgery) assessments, including clinical ophthalmic evaluations, MRI, and pupillometry. Healthy controls underwent a single baseline assessment, consisting of ophthalmic evaluations and pupillometry.

### Clinical evaluation and imaging

#### Demographics and medical history

The data collected included age, gender, ethnicity, medical history (e.g., diabetes, systemic conditions), ocular history, medication use, and smoking status.

#### Ophthalmic assessments

All participants underwent a comprehensive ophthalmic evaluation including measurement of best-corrected visual acuity (BCVA) using a LogMAR chart, retinal nerve fiber layer imaging using high-definition OCT, and visual field testing using standard automated perimetry (Humphrey Visual Field (HVF) analyzer).

#### MRI assessment

Pre-operative and post-operative MRI scans were obtained for all patients except two who lacked pre-operative scans due to assessments at another hospital. Fine-cut coronal T2-weighted MRI scans were used to assess the upward displacement of the optic chiasm and optic chiasm thickness. All MRI evaluations were conducted by a neurosurgeon, who extracted optic chiasm displacement and thickness measurements for analysis.

### Handheld chromatic pupillometry

Pupillometry testing was conducted using a standardized 1-minute protocol with a custom-built handheld chromatic pupillometer(23). The test was conducted monocularly in a dark room (<1 lux) with alternating blue- and red-light stimulations and interleaved dark phases (Figure 1A), following identical stimulus parameters, recording setup, and analysis procedures established in prior studies using the same HCP device(16–18,24). Please see supplementary materials for more details about the HCP protocol.

**Figure 1.**
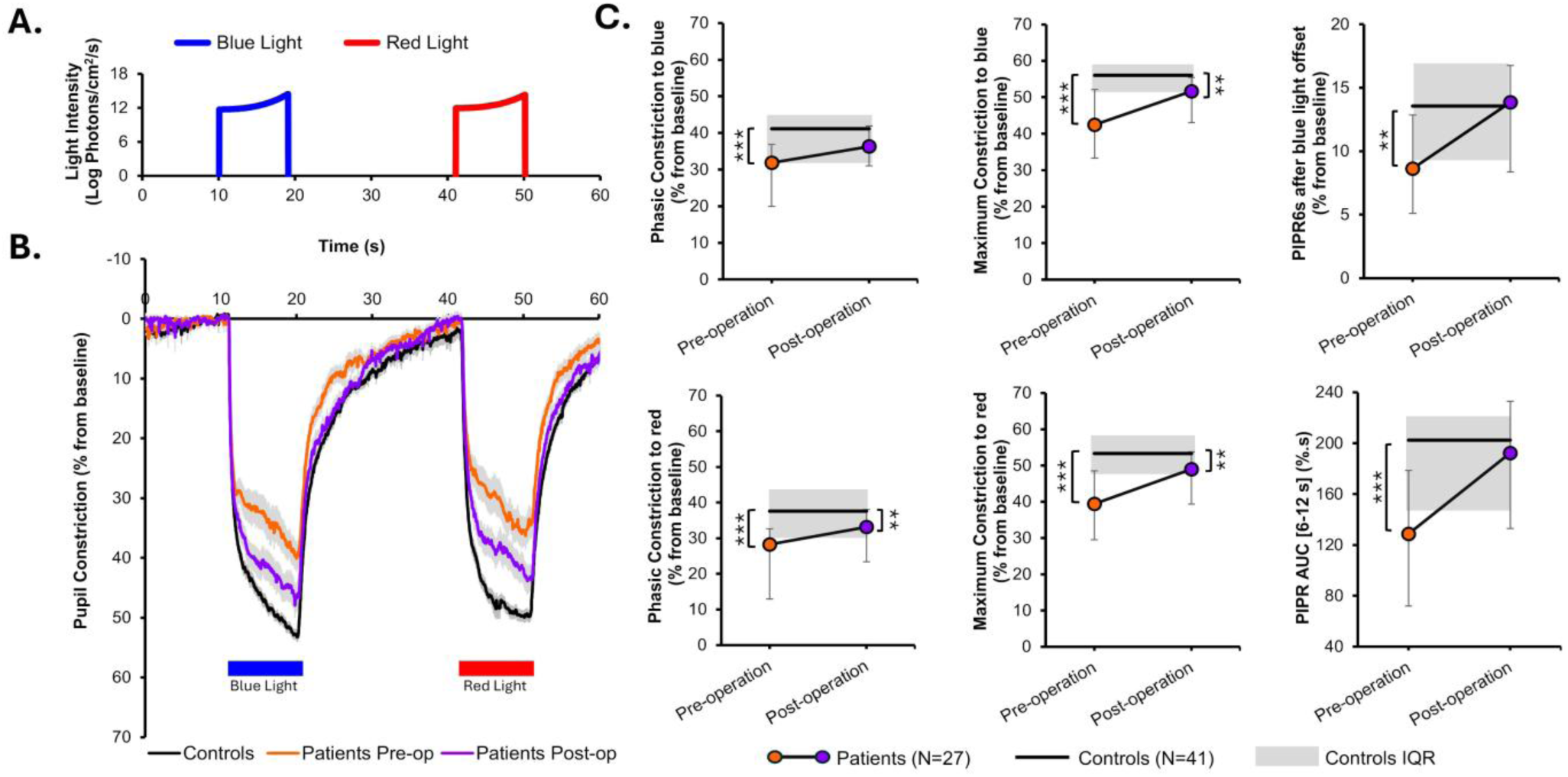
Average baseline-adjusted pupillary light responses in patients with pituitary adenoma (n=27) and controls (n=41). **A.** Pupillometry light protocol. **B.** Mean pupillary responses to blue and red light in patients with pituitary adenoma (n=27) before and after surgery and controls. Data are plotted as average ± SE. **C.** Differences in the median of the main pupillometric features in patients with pituitary adenoma and controls. Statistical comparisons between groups were performed using a Mann-Whitney U test. Error bars represent SE and connecting lines illustrate individual changes. **p<0.01; ***p<0.001. **Abbreviations:** AUC, area under the curve; IQR, interquartile range; PIPR, post-illumination pupillary response.

### Data analysis and statistics

Pupil radius measurements were processed using a semiautomated algorithm for blink artefact removal and expressed as a percentage change from baseline pupil size. Sixteen pupillometric features were extracted from individual blink-free traces for further analysis, among which we have phasic constriction to blue (Phasic-Blue) and red (Phasic-Red) light, maximum constriction to blue (Max-Blue) and red (Max-Red) light, post illuminance pupillary response 6 (PIPR6s) and 12 seconds after light offset (PIPR12s), PIPR area under the curve (PIPR AUC 0–12s), redilation slope 1.7s following light offset (PIPR>1.7 slope).

Optic chiasm displacement and thickness were measured from fine-cut coronal T2-weighted MRI scans to the nearest 0.1 mm. The optic chiasm was identified by tracing bilateral optic nerves backwards from the retina, through the intraconal portion into the optic canal, and then subsequently superior to the pituitary tumour. Three measurements were taken for each parameter on each MRI, and their average was used for analysis.

All pupillometric, MRI, and ophthalmic features are reported as median (IQR) and were compared between controls and PA patients using the Mann-Whitney U test. Comparisons of pre- and post-operative outcomes in PA patients were performed using the Wilcoxon signed-rank test. Associations between structural and functional outcomes were assessed using Spearman’s rank correlation coefficient. Demographic data are presented as median (Interquartile range (IQR)) or number (%) and were compared between controls and PA groups using either the Mann-Whitney U test or χ² test, as appropriate. Recovery was defined as the point at which a patient’s pupillometric or ophthalmic features returned within the 95% confidence interval (CI) of control values. All statistical analyses were conducted using SPSS Statistics v26 (IBM, United States).

## RESULTS

### Demographics and clinical characteristics of participants

Data from 41 healthy controls (median age = 55.4 years (IQR = 23.4), 46.3% males, 97.6% ethnic Chinese) and 27 PA patients (53.6 (13.7) years, 40.7% males, 74.1% ethnic Chinese) were analyzed. Groups were age-matched (p=0.72). The control group had a higher proportion of Chinese participants (p=0.03). Other baseline clinical characteristics, including diabetes, cataract history, and prior posterior chamber intraocular lens (PCIOL) implantation, were similar between groups (all p>0.05). A detailed breakdown of demographic and clinical characteristics for the total sample is presented in Table 1, while subgroup characteristics are provided in Supplementary Tables 1, 2, and 3.

**Table 1.**
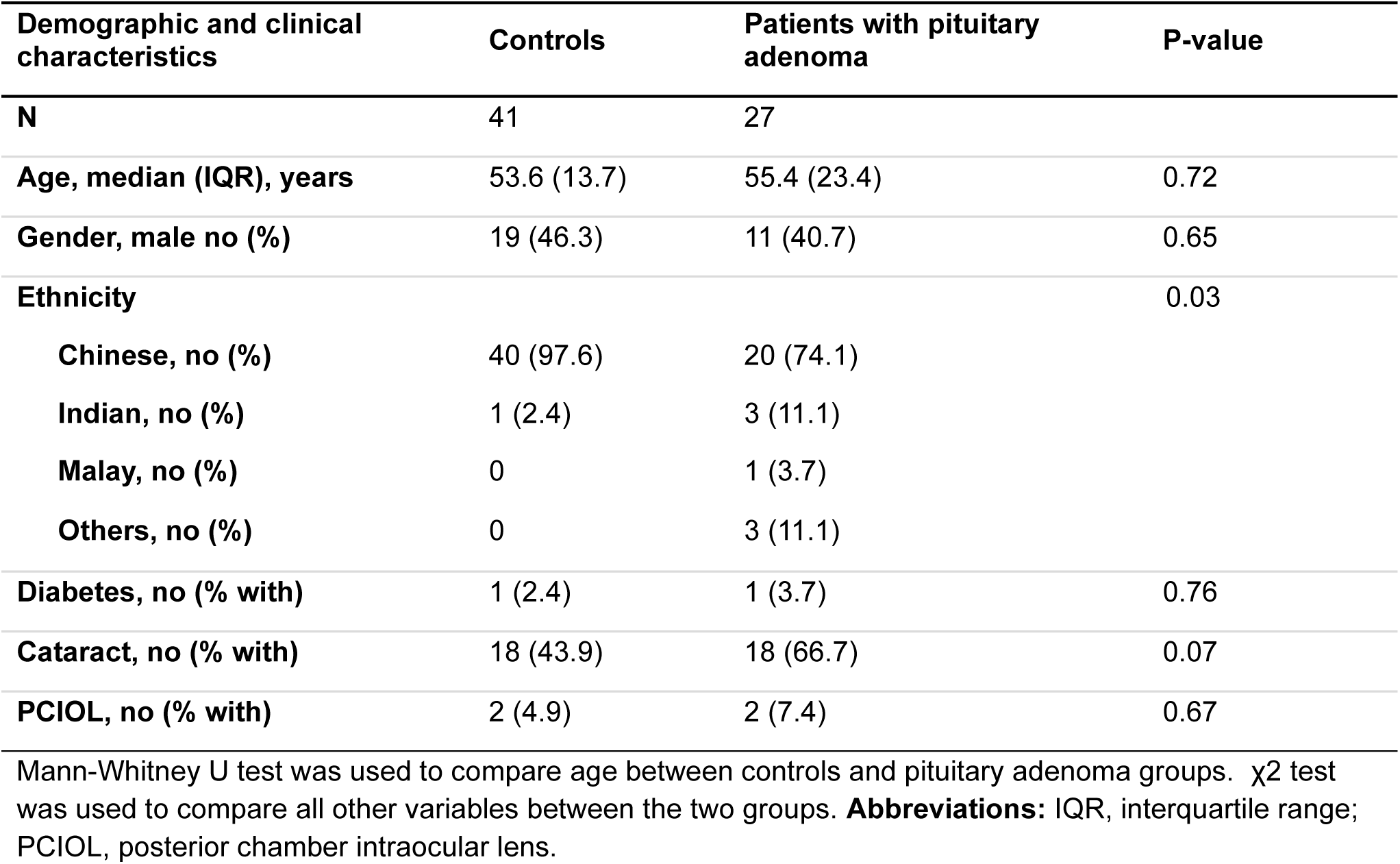
Demographics and clinical characteristics of patients with pituitary adenoma (n=27) and healthy controls (n=41)

### Pre-operative pupillary light responses in patients with pituitary adenoma

Prior to TSS, patients with PA exhibited significant differences in the baseline-adjusted pupillary responses to both blue- and red-light stimuli compared to healthy controls (Figure 1B). Patients with PA demonstrated prolonged constriction latency (Controls: 0.35 s (IQR: 0.17), Patients: 0.48(0.31) s for blue light; Controls: 0.47(0.21) s, Patients: 0.58(0.82) s for red light), reduced phasic constriction (Controls: 41.1(12.6)%, Patients: 31.9(16.9)% for blue; Controls: 37.6(12.7)%, Patients: 28.3(19.7)% for red light) , and lower maximum constriction (Controls: 56.0(6.8)%, Patients (42.4(18.8)% for blue light; Controls: 53.3(9.8)%, Patients: 39.4(18.9)% for red light) (all p<0.001 for both stimuli). Pupillary redilation following blue-light exposure was also impaired, with diminished PIPR at 6 seconds (Controls: 13.6(7.5)%, Patients: 8.6(7.8)%, p=0.001) and 12 seconds (Controls: 6.4(8.3)%, Patients: 2.8(5.3)%, p=0.04) after blue light offset, a significantly reduced PIPR AUC 0–12s (Controls: 202.5 (79.4)%.s, Patients: 128.6(106.8)%.s, p<0.001) compared to controls (Figure 1C), and a smaller PIPR>1.7 slope (Controls: -1.0(0.6)%/s, Patients: -0.7(0.5)%/s, p=0.001) (Supplementary Figure 2; Supplementary Table 4).

### Recovery in pupillometric features following surgical decompression

Following TSS, constriction latency improved but remained higher than controls (Controls: 0.35(0.17) s, Patients: 0.40(0.26) s for blue light; Controls: 0.47(0.21) s, Patients: 0.56(0.30) s for red), though this difference was not statistically significant. Maximum constriction to both stimuli increased post- operatively but did not return to control levels (Controls: 56.0(6.8)%, Patients: 51.6(12.4)%, p=0.007 for blue; Controls: 53.3(9.8)%, Patients: 49.0(14.4)%, p=0.01 for red). In contrast, blue PIPR6s (Controls: 13.6(7.5)%; Patients: 13.9(8.4)%) and PIPR AUC 0–12s (Controls: 202.5(79.4)%.s, Patients: 192.3(100.1)%.s) normalized post-surgery, reaching control values (Figure 1C; Supplementary Table 4).

### Associations between structural and functional recovery following surgery

Paired comparisons of MRI features before and after surgery showed significant reductions in the upward displacement of the optic chiasm (UDOC) (Pre-op: 6.3(3.8) mm, Post-op: 0.8(1.6) mm, p<0.001, Figure 2A) and an increase in optic chiasm thickness (OpCT) (Pre-op: 1.1(0.5) mm, Post-op: 2.4(0.6)mm, p < 0.001, Figure 2B), indicating effective structural decompression. OCT data from 17 patients showed no difference in RNFL thickness pre- (88(9) µm) versus post-surgery (86(5) µm; p>0.05). However, RNFL thickness remained significantly thinner compared with controls (96(5) µm; p<0.05 for both pre- and post-surgery comparisons). Comparatively, functional recovery was also noticed post-surgery. BCVA, as well as visual field outcomes such as the visual field index (VFI), visual field mean deviation (VFMD) and pattern standard deviation (PSD) improved post-surgery (all p<0.05; Figure 2C-F) but did not reach control levels (Supplementary Table 1, Supplementary Figure 3).

**Figure 2.**
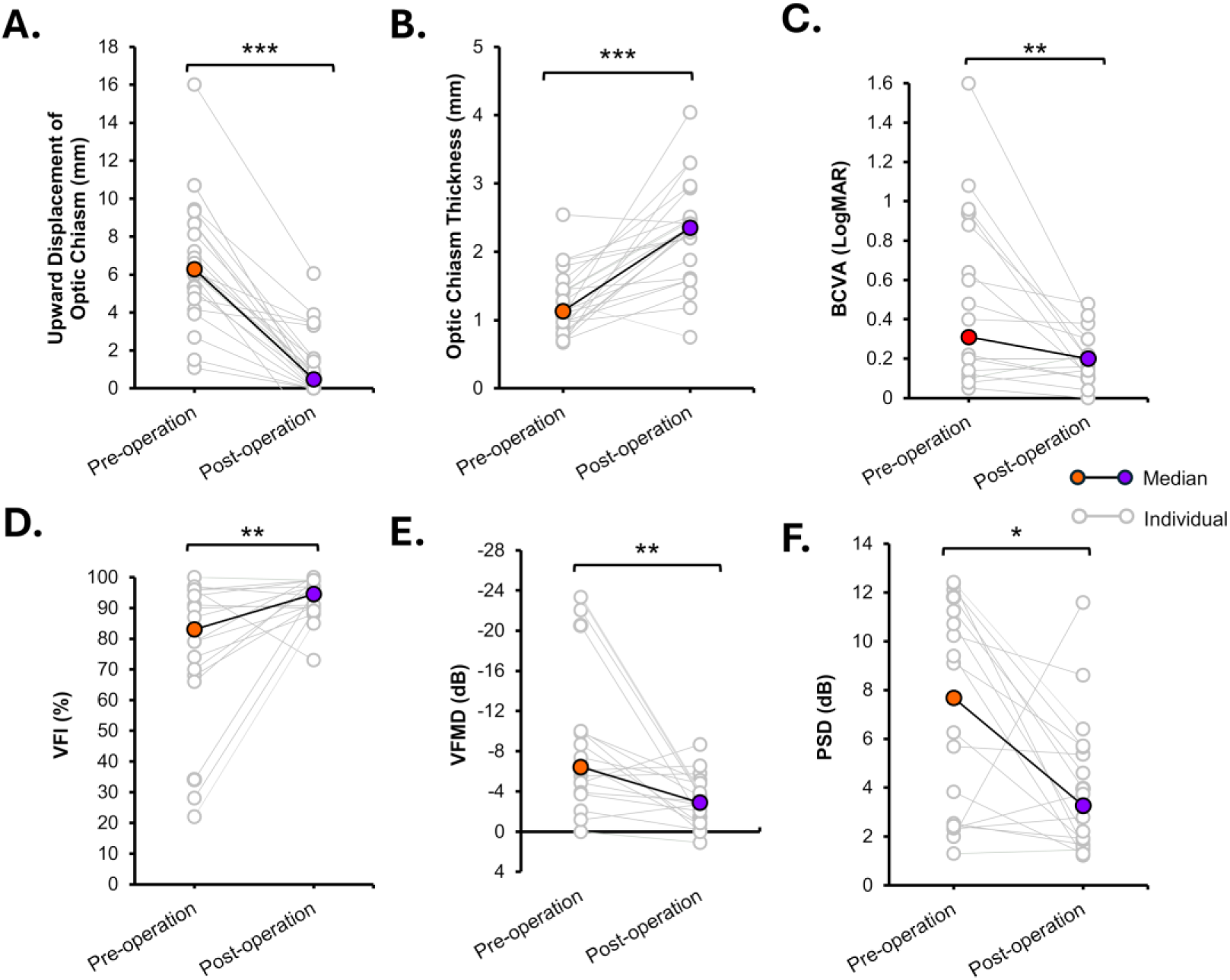
Paired comparison of pre-operative and post-operative patient outcomes. **A-B.** Structural MRI outcomes (n=25) **C**. Best-corrected visual acuity (BCVA) (n=18) **D-F.** Humphrey visual field (HVF) outcomes (n=18). *p<0.05; ** p<0.01; ***p<0.001. **Abbreviations:** BCVA, best-corrected visual acuity; LogMAR, logarithm of minimum angle of resolution; PCIOL, posterior chamber intra-ocular lens; PSD, pattern SD; VFI, visual field index; VFMD, visual field mean deviation.

Spearman correlation analysis revealed a significant negative correlation between UDOC and VFI (ρ= -0.62, p<0.001; Figure 3A) as well as UDOC and Max-Red (ρ= -0.47, p=0.004; Figure 3B). Conversely, UDOC was positively correlated with PIPR>1.7 slope (ρ= 0.41, p=0.01; Figure 3C). In contrast, changes in UDOC were not correlated with changes in BCVA (ρ= 0.30, p=0.08). Similarly, Max-Red and BCVA were not correlated (ρ= -0.11, p=0.51), while Max-Red and VFI showed a marginal correlation that did not reach statistical significance (ρ = 0.32, p=0.06). A comprehensive overview of all correlations is provided in the supplementary material (Supplementary Figure 4).

**Figure 3.**
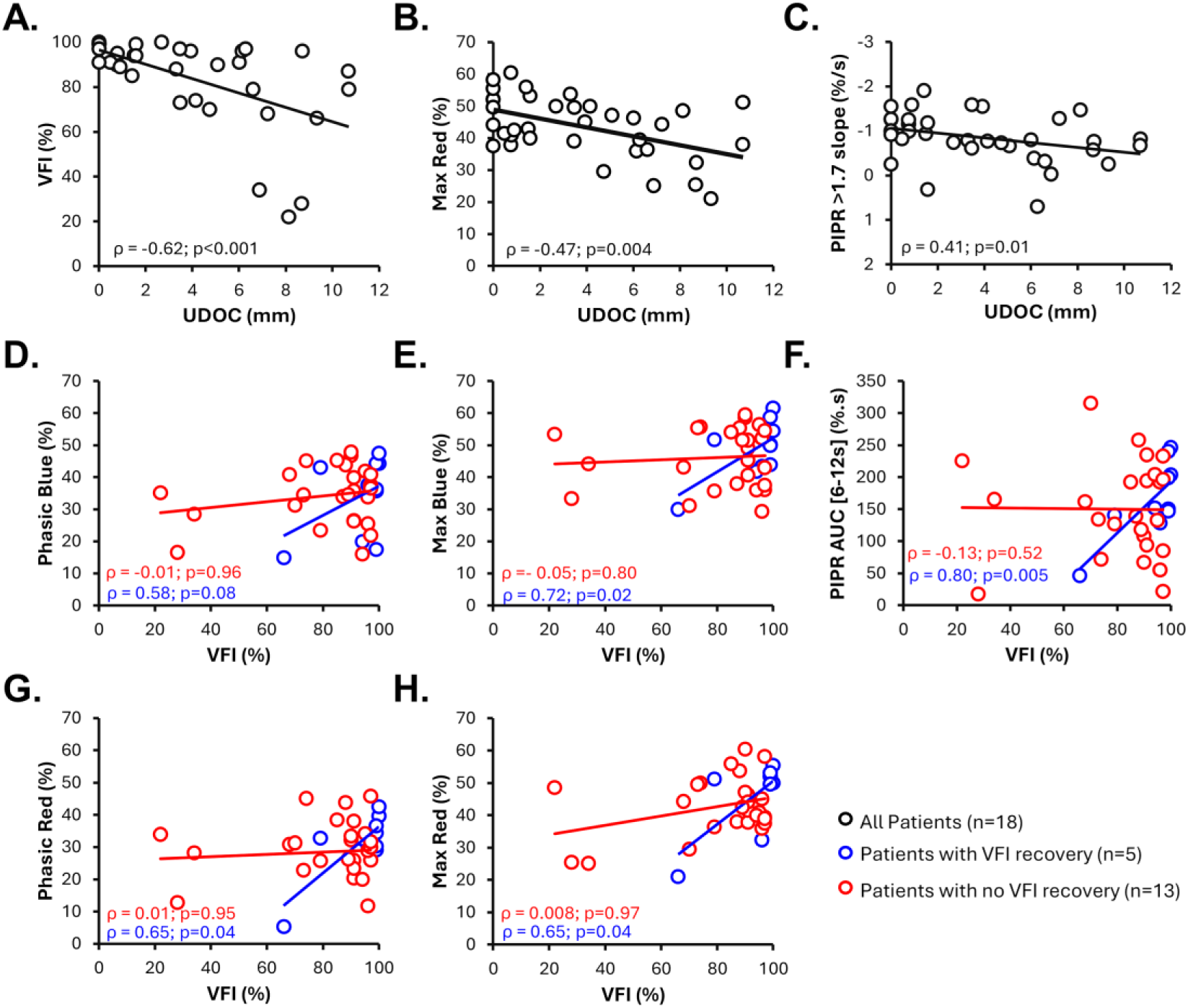
Scatter plots showing Spearman correlations. Each point represents a patient’s pre- or post-operative measurement. **A–C**: Correlations between structural (UDOC) and functional (VFI, Max Red, PIPR>1.7 slope) outcomes. **D–H**: Correlations between functional ocular (VFI) and functional pupillometric outcomes. **Abbreviations:** AUC, area under the curve; BCVA, best-corrected visual acuity; Max Red, maximum constriction to red light; PIPR>1.7 slope, post-illumination pupillary response at >1.7 slope; UDOC, upward displacement of the optic chiasm; VFI, visual field index.

Subgroup analyses classified patients based on whether their VFI fell within the 95% CI of healthy controls, with those meeting this threshold considered to have achieved full recovery. Although no pupillometric feature was significantly correlated with VFI in all 18 participants, stratified analysis revealed that several pupillometric features, including Phasic-Red (Figure 3G), Max-Blue and Max-Red, and PIPR AUC 0-12s, were significantly correlated with VFI in patients who achieved full recovery (n=5; all p<0.05), but not in those who did not (n=13; all p>0.5) (Figure 3E-H).

### Visual field recovery and pupillometric features

Patients with and without visual field recovery post-surgery exhibited distinct trajectories in PLR parameters. Patients who achieved visual field recovery demonstrated post-surgical phasic constriction and maximum constriction at control levels (p>0.05 for both lights; Figure 4). In contrast, patients without visual field recovery exhibited persistent deficits, including a 5.9% reduction in Phasic-Red (p=0.03) and reduction in Max-Blue (4.4%) and Max-Red (10.8%) (both p=0.01) compared to controls (Supplementary Figure 5).

**Figure 4.**
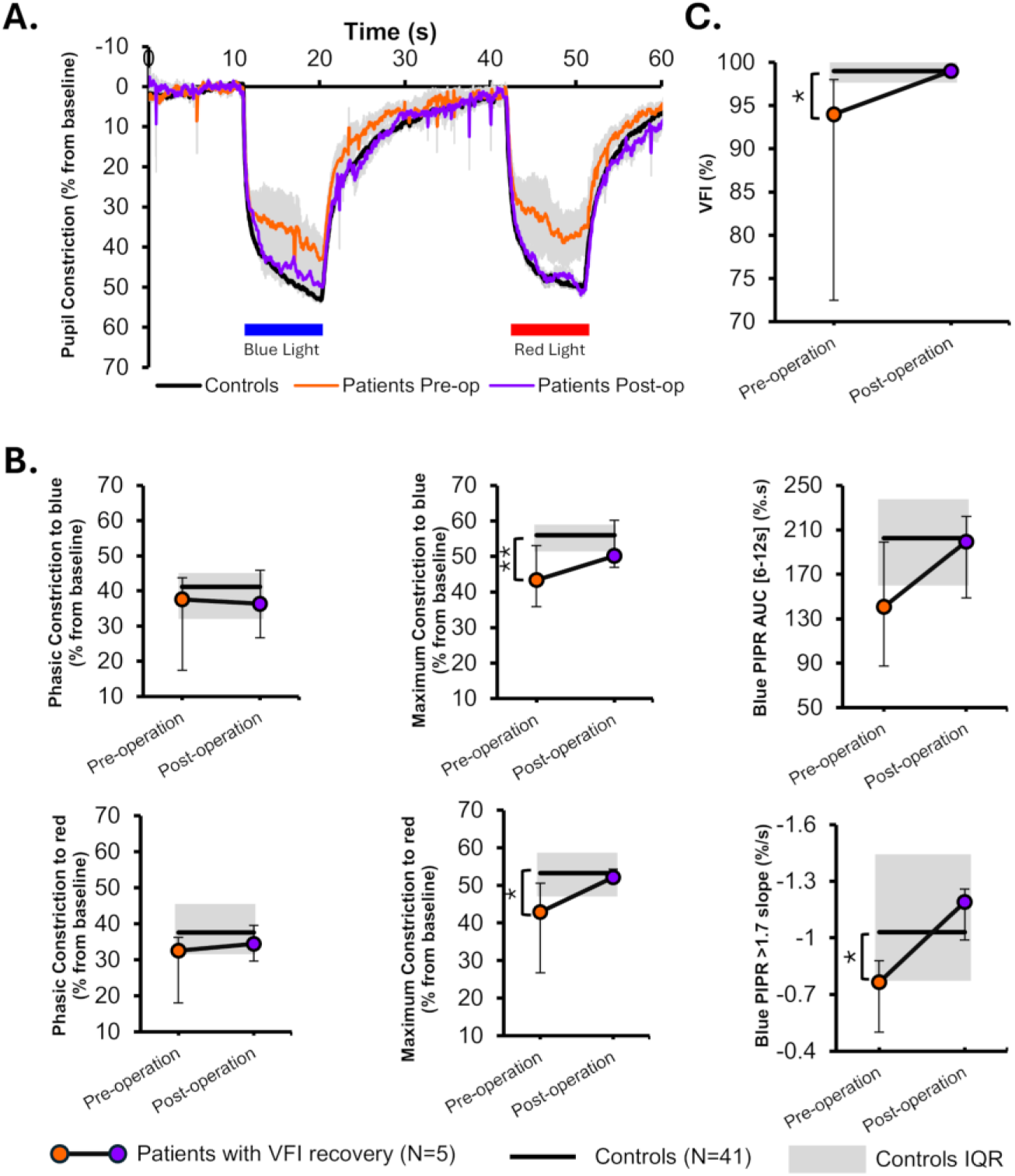
PLR and comparisons of pupillometric parameters in patients with VFI recovery. **A.** Mean pupil response traces to blue and red-light stimuli, in patients with Pituitary Adenoma with VFI recovery (n=5) before and after surgery and controls. **B.** Differences in main pupillometric features between patients pre- and post-operation and controls. **C.** Difference in VFI in patients with PA before and after surgery. *p<0.05; ** p<0.01. **Abbreviations:** AUC, area under the curve; IQR, interquartile range; VFI, Visual Field Index; PIPR, post-illumination pupillary response; PLR, pupillary light response.

Despite these group differences in phasic and maximum constriction, post-surgery pupillometric redilation features (i.e., blue PIPR6s, blue PIPR12s, PIPR AUC 0-12s, and PIPR>1.7 slope) were comparable to controls in both recovered and non-recovered patients (all p>0.05).

### Enhanced detection of pupillary deficits using chromatic pupillometry

Pre-operatively, clinical examination revealed no relative afferent pupillary defect (RAPD) in 11 of 18 patients; however, all of these patients exhibited pupillometric abnormalities when assessed with direct monocular HCP. Following surgery, 16 patients demonstrated no clinical RAPD. Of these, 9 continued to show residual pupillometric defects on HCP. Among the 16 patients without clinical RAPD post-operatively, 5 achieved complete visual field recovery, and 4 of these patients also demonstrated full recovery on HCP.

### Individual variability in recovery trajectories

Individual patient data demonstrated a range of recovery outcomes, with some patients achieving near-normal PLR responses post-operatively, while others exhibited persistent deficits.

### Case 1: Complete recovery

Patient 1 presented with a large PA compressing the optic chiasm, as evidenced by pre-operative MRI (9.33 mm displacement) and severe visual field deficits. Pre-operative pupillometry revealed abnormal PLR responses to both blue and red light. Post-surgery, MRI confirmed significant decompression of the optic chiasm (1.57 mm), and visual field testing showed substantial improvement. Pupillometry findings normalized across all features, aligning with improvements in visual field outcomes. No RNFL changes were noticed (RNFL-Pre: 82 μm, RNFL-Post: 81 μm). This case highlights complete recovery across structural (MRI), functional (pupillometry), and visual (HVF) domains, emphasizing the potential for full restoration of visual pathways following TSS in cases with significant decompression (Figure 5).

**Figure 5.**
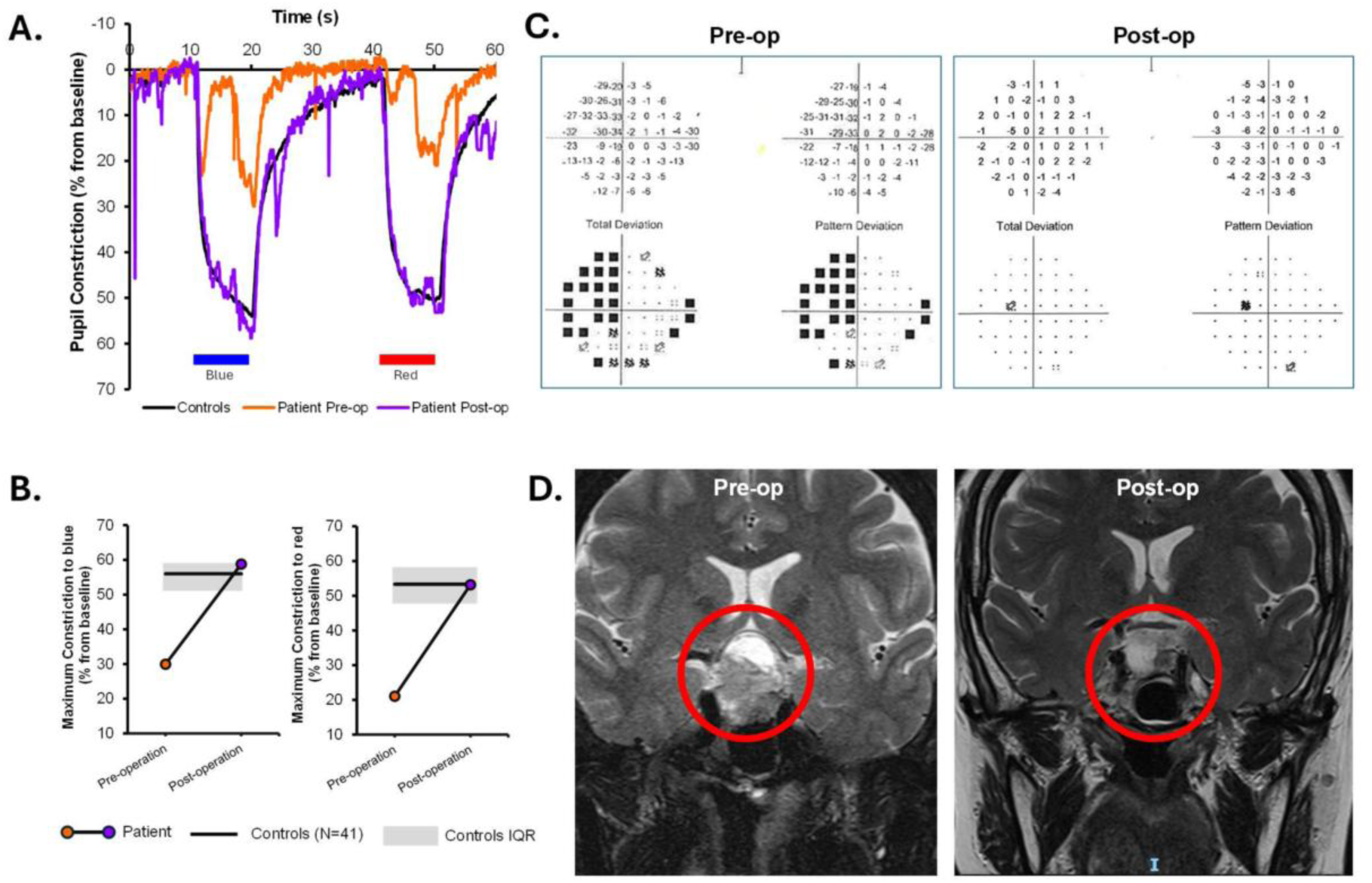
Patient 1 multimodal assessment of visual and pupillary function pre- and post-surgery. **A.** Pupil response traces. **B.** Maximum pupillary constriction to blue and red light pre- and post- operation **C.** Visual field assessment **D.** Tumor resection onT2-weighted MRI images before and after surgery.

### Case 2: Partial recovery

Patient 2, with a large PA compressing the optic chiasm, presented with pre-operative visual field deficits. Pre-operative MRI revealed 6.6 mm upward displacement of the optic chiasm. Pupillometric assessment showed severe deficits in response to both blue and red-light stimuli. Post-surgical MRI confirmed effective decompression of the optic chiasm (0 mm), and visual field testing showed improvement but not complete recovery. Post-operative pupillometry indicated recovery in pupillary responses to blue light and improvement in red light-mediated responses, though residual deficits persisted in response to red-light in particular. No RNFL recovery was noticed (RNFL-Pre: 85 μm, RNFL-Post: 82 μm). This case demonstrates partial recovery for functional visual domains, with residual deficits in cone-mediated responses indicating the complexity of functional restoration even with effective structural decompression (Figure 6).

**Figure 6.**
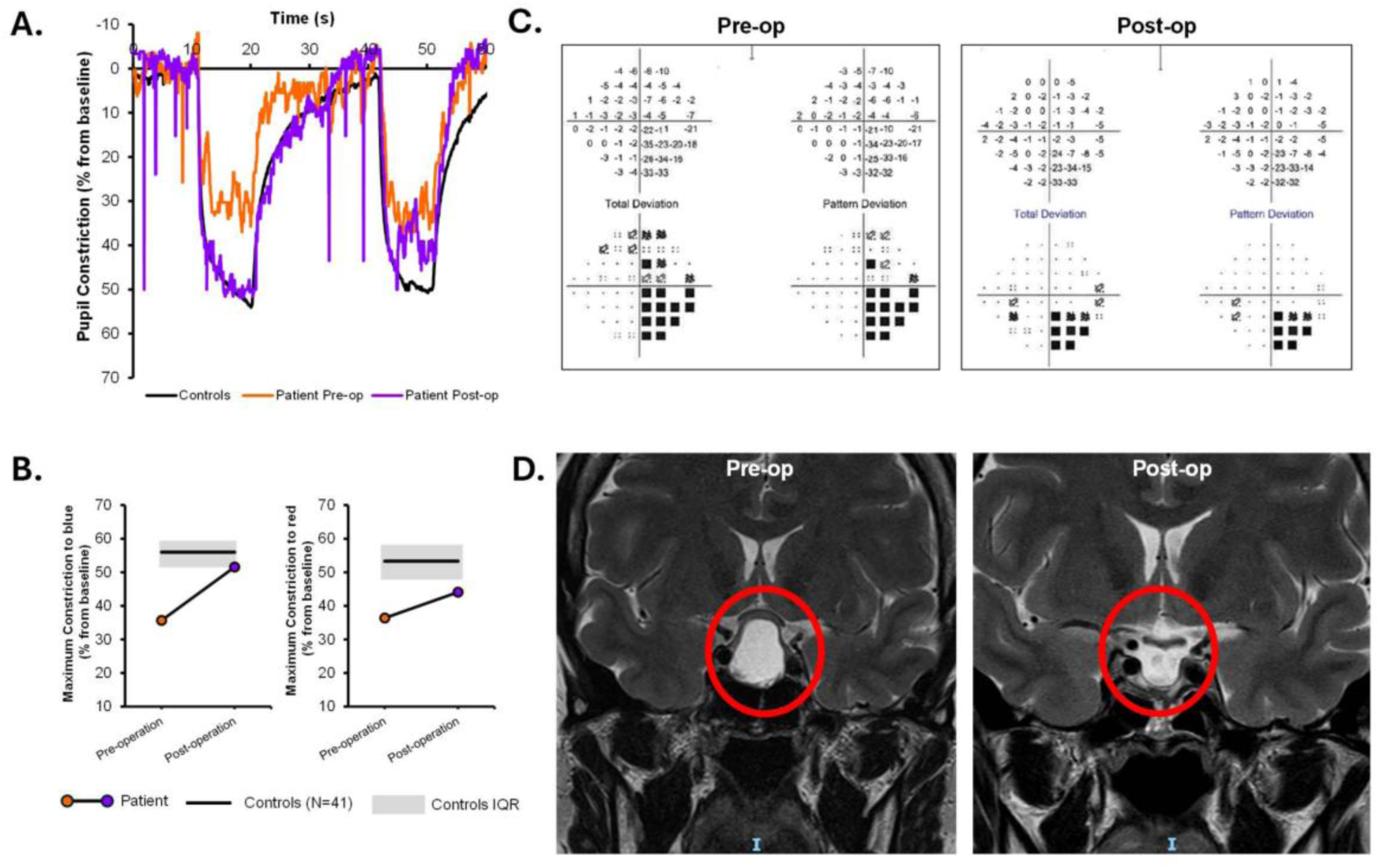
Patient 2 multimodal assessment of visual and pupillary function pre- and post-surgery. **A.** Pupil response traces. **B.** Maximum pupillary constriction to blue and red light pre- and post-operation, **C.** Visual field. **D.** Tumor resection on T2-weighted MRI images before and after surgery.

### Case 3: Limited recovery

Patient 3 presented with a PA causing marked optic chiasm compression, as confirmed by pre- operative MRI (4.73 mm displacement). The patient reported visual field loss, with pre-operative visual field testing revealing extensive deficits. Pre-operative pupillometric assessments showed alterations in response to both blue and red-light stimuli. Post-operative MRI demonstrated partial decompression of the optic chiasm (3.46 mm), but residual compression was still evident. Visual field testing showed recovery, with residual defects persisting. Post-operative pupillometry revealed slight improvement in response to blue light; however, red light-mediated responses remained significantly impaired. No RNFL recovery was noticed (RNFL-Pre: 89 μm, RNFL-Post: 86 μm). This case highlights the variability in outcomes and the potential for residual deficits following TSS (Figure 7).

**Figure 7.**
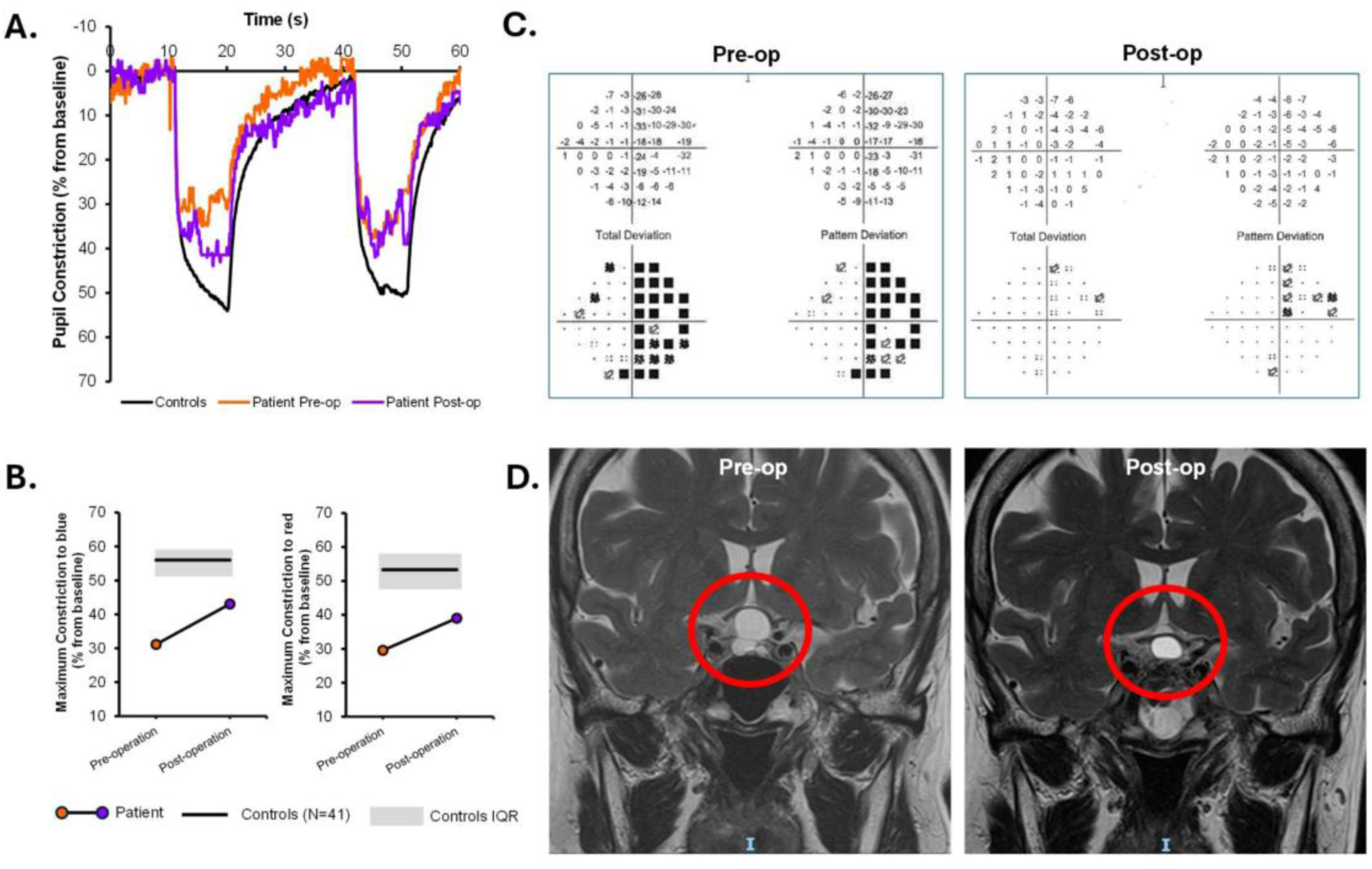
Patient 3 multimodal assessment of visual and pupillary function pre- and post-surgery. **A.** Pupil response traces. **B.** Maximum pupillary constriction to blue and red light pre- and post-operation. **C.** Visual field assessment. **D.** Residual tumor on T2-weighted MRI images before and after surgery.

## DISCUSSION

This study evaluated visual and non-visual pupillometric functions in PA before and after TSS. HCP proved to be a feasible, objective, and non-invasive tool for assessing the functional disruption and restoration of the visual and non-visual pathways. Notably, irrespective of the visual outcomes, pituitary adenoma surgery resulted in consistent improvements in PLR, aligning with structural and functional recovery. Importantly, melanopsin-driven responses normalized after surgery even in individuals without visual field recovery, indicating that non-visual pathways may restore independently of vision.

In line with previous studies(12,13), PA patients exhibited significant pre-operative impairments in pupillary responses to blue and red-light stimuli, suggesting dysfunction in both intrinsic (melanopsin-driven) and extrinsic (cone-mediated) pathways due to optic chiasm compression. Pupillometric dysfunction in PA, align with previous findings in glaucoma(16,25) and diabetic retinopathy(18), diseases affecting ipRGC integrity. The optic chiasm compression in PA can lead to retrograde degeneration of retinal ganglion cells(26), presented on OCT as thinning of the retinal nerve fiber layer (RNFL) and ganglion cell layer (GCL) in affected patients (27).

Post-operative MRI confirmed effective decompression of the optic chiasm, with reduced chiasmal displacement and increased chiasm thickness. However, functional outcomes varied considerably among patients, showing that structural restoration does not always equate too complete functional recovery. This variability is consistent with prior studies demonstrating that post-surgical visual recovery is influenced by factors such as the degree of pre-operative damage, duration of symptoms, and individual neuroplasticity(28–30). Although some patients exhibited near-normalization of PLR, others showed persistent deficits despite apparent structural improvements on MRI. These findings emphasize the need for a functional assessment of visual pathways in addition to structural imaging, as anatomical normalization alone may not fully capture the extent of neuro-ophthalmic recovery. Additionally, VFI correlated well with structural features and showed a trend for correlation with pupillometry, supporting its use as a more relevant functional outcome in PA compared to visual acuity.

A key finding of this study is the differential recovery observed between melanopsin-driven and cone-mediated responses. While PIPR to blue light (i.e. PIPR6s, PIPR AUC 0-12s, PIPR >1.7 slope), primarily mediated by melanopsin,(31), and reduced in patients with glaucoma(16), fully recovered, maximum and phasic constriction, predominantly cone-mediated, remained impaired. These findings suggest that intrinsic ipRGC pathways may be more resilient to compressive damage and capable of greater functional recovery than extrinsic cone-mediated pathways(32). This differential recovery may reflect both anatomical and molecular properties of ipRGC subtypes. M1 ipRGCs, rich in melanopsin, show strong intrinsic photosensitivity and are known to resist optic nerve injury more effectively than other subtypes(33). In contrast, M2 ipRGCs receive substantial extrinsic input from cones and bipolar cells(34) and appear more vulnerable to structural damage(35). Both M1 and M2 project to the olivary pretectal nucleus and contribute to the pupillary light reflex(36), but their differential resilience may account for the partial restoration observed in our study. While recovery of PIPR likely reflects the robustness of M1 ipRGCs, the persistence of deficits in maximum and phasic constriction may result from residual dysfunction in cone pathways, disrupted synaptic integration, or damage to more vulnerable ipRGC subtypes such as M2. Whether the selective ipRGC recovery observed represents true neuroregeneration or compensatory mechanisms remain unclear and warrants further investigation using approaches that can differentiate ipRGC subtypes and their specific projections. Conversely, direct cone dysfunction in PA is not well established in the literature, it is possible that cone pathways are secondarily affected by RGC degeneration. Experimental models have shown that long-term ischemic damage can extend beyond inner retinal layers to affect outer retinal structures, including cone pathways(37). Moreover, ipRGC degeneration could lead to neurovascular alterations that indirectly impact cone function, and chronic compression has been shown to trigger retinal remodeling, involving neuronal cell death and synaptic rewiring(38)

Correlations between visual field recovery and PLR improvement further reinforces the potential role of pupillometry as a biomarker for neuro-ophthalmic recovery following TSS. Patients with greater VFI recovery show enhanced PLR, particularly in phasic and maximum constriction. Given that perimetry relies on subjective patient responses and requires sustained attention(8), pupillometry offers an objective, and quantifiable alternative for assessing visual function. Furthermore, compared to clinical RAPD assessment, direct HCP seems to provide a more sensitive approach for detecting and monitoring functional deficits in patients with PA.

This study is the largest to date investigating pupillometry in pituitary adenomas, yet some limitations should be acknowledged. First, the sample size remains relatively small for predictive modeling and larger cohorts are needed to strengthen statistical power and validate pupillometric predictors of visual recovery. Second, some MRI, visual field, and OCT data were missing due to logistical issues, reducing the number of complete cases and leading to stratification. Third, the ∼180° field chromatic pupillometry protocol may have missed localized retinal dysfunction corresponding to compressed visual field regions, suggesting that spatially targeted approaches could enhance topographic assessment. Fourth, pupillary responses measured in our study may not strictly reflect the laboratory-defined PIPR, which typically requires pharmacological dilation, brief and intense light stimuli, and shorter exposure durations (31,39). Nevertheless, similar pupillometric features, obtained using the same HCP device, have previously been shown to be reduced in glaucoma patients, a population known to exhibit impaired intrinsically photosensitive retinal ganglion cell (ipRGC) function(16). Finally, our protocol was limited to a short follow-up period of ∼12 weeks, which may have missed long-term functional changes following surgery.

In conclusion, our findings demonstrate that although TSS for PAs leads to significant structural decompression of the optic chiasm, functional recovery of the PLR remains incomplete in certain cases. Notably, ipRGC-mediated responses, measured by the PIPR to blue light, fully recover post-operatively, whereas cone-mediated pathways, assessed by maximum and phasic constriction, show persistent impairments. The preservation and restoration of melanopsin-driven ipRGC responses are especially notable, as these pathways play a critical role in circadian entrainment, sleep regulation, and other systemic processes(40). HCP thus provides a sensitive, objective, and non-invasive biomarker of recovery, correlating with visual field outcomes and offering clinical value where conventional testing is limited. Integrating HCP into routine clinical workflows could provide a more holistic understanding of surgical outcomes, supporting both neuro-ophthalmic and systemic health monitoring in patients with pituitary adenoma. Future research should explore predictive models based on pre-operative pupillometric features and evaluate long-term systemic consequences, ultimately guiding individualized management and improving quality of life in patients with pituitary adenoma.

## Acknowledgments

The authors thank the clinical research coordinators for their valuable assistance in patient recruitment and data collection and extend their gratitude to all participants for their time and contribution to this study. Part of this work was previously presented at the 2025 Annual Meeting of the Association for Research in Vision and Ophthalmology (ARVO): Mahfoud D, Ang J, Ang BT, Milea D, Najjar RP. Pupillometric features of functional ocular recovery following transsphenoidal surgery in patients with pituitary adenoma. Invest Ophthalmol Vis Sci. 2025;66(8):3223.

## Contributors

Concept and design: RPN, DMI, ABTI. Data acquisition and research execution: RPN, JA. Analysis and interpretation: DM, JA, DMI, ABTI, MEN, and RPN. Manuscript preparation: DM, JA, ABTI, DMI, MEN and RPN.

## Patient consent for publication

Consent obtained directly from patient(s) Ethics approval. The study was approved by the SingHealth Centralised Institutional Review Board (CIRB (2018/3233)).

## Data availability statement

The de-identified datasets and the study protocol can be made available from the corresponding author upon reasonable request.

