## Supplementary Material for "Recovery in pupillometric non-visual functions following chiasmal decompression in pituitary adenoma"

***Corresponding Author**

### This supplementary material document includes:

**Supplementary Methods:**

1. **Eligibility criteria**
2. **Handheld chromatic pupillometry**

**Supplementary Tables:**

1. **Supplementary Table 1.** Demographics and clinical characteristics of patients with pituitary adenoma with complete dataset (n=18) and healthy controls (n=41)
2. **Supplementary Table 2.** Demographics and clinical characteristics of patients with pituitary adenoma with visual field recovery (n=5) and healthy controls (n=41)
3. **Supplementary Table 3.** Demographics and clinical characteristics of patients with pituitary adenoma with no visual field recovery (n=13) and healthy controls (n=41)
4. **Supplementary Table 4.** Pupillometric features in patients with pituitary adenoma pre- and post- surgery (n=27) compared to healthy controls (n=41)

**Supplementary Figures:**

1. **Supplementary Figure 1.** Flowchart of enrolment and exclusion of patients and data
2. **Supplementary Figure 2.** Pupillary light reflex (PLR) parameters pre- and post-surgery (n=27) compared to controls (n=41)
3. **Supplementary Figure 3.** Ophthalmic assessments pre- and post- surgery (n=18) compared to controls (n=41)
4. **Supplementary Figure 4.** Heatmap illustrating the spearman correlation coefficients between all measured variables
5. **Supplementary Figure 5.** Pupillary light response and comparisons of pupillometric parameters in patients without full VFI recovery

### Supplementary Methods

#### Eligibility Criteria

Patients were eligible if they had a PA causing radiological optic chiasm indentation or distortion and were scheduled for surgical resection. Patients and controls were aged 21 years or older. Exclusion included a history of retinopathy and/or optic neuropathy, raised intraocular pressure, or glaucoma in either eye, refractive error exceeding ± 6.0 DSph or ± 3.0 DCyl, previous intraocular surgery (except uncomplicated cataract procedures), cataract severity worse than NS2+, participants on any drugs that may affect pupillary size or responses, or having conditions affecting afferent or efferent pupillary pathways, and participants with previous trauma to the eyes or previous intraocular inflammation.

#### Handheld chromatic pupillometry

Pupillometry testing was conducted using a standardized 1-minute protocol with a custom-built handheld chromatic pupillometer in a darkened room (<1 lux). The device, designed for monocular use, included a silicone rubber eye cup for comfortable positioning over the study eye and ensured light isolation, with the fellow eye covered by the participant's hand. The protocol involved five consecutive phases: 10 seconds of darkness for baseline pupil size measurement, 9 seconds of exponentially increasing blue light stimulation (11.7–14.4 Log photons/cm²/s; $\lambda max$ = 469 nm, FWHM = 33 nm), 22 seconds of darkness for pupillary redilation, 9 seconds of exponentially increasing red light stimulation (11.9–14.3 Log photons/cm²/s; $\lambda max$ = 640 nm, FWHM = 17 nm), and a final 10 seconds of darkness to assess redilation. Participants fixated on a central dim red zone (<0.1 lux) within the device, and any issues with fixation or excessive blinking prompted a repeat of the test. An infrared camera positioned at ~60° below the lower eyelid recorded horizontal pupil size changes at a frame rate of 30 fps, while light stimuli were calibrated to stimulate 180° of the visual field.

### Supplementary Tables

**Supplementary Table 1. Demographics and clinical characteristics of patients with pituitary adenoma with complete dataset (n=18) and healthy controls (n=41)**

| **Demographic and clinical characteristics** | **Controls** | **Patients with pituitary adenoma** | | **P-value** |
|---|---|---|---|---|
| **N** | 41 | 18 | | |
| **Age, median (IQR), years** | 53.6 (13.7) | 57.4 (22.4) | | 0.35 |
| **Gender, male no (%)** | 19 (46.3) | 7 (38.9) | | 0.60 |
| **Ethnicity** | | | | |
| **Chinese, no (%)** | 40 (97.6) | 15 (83.3) | | 0.08 |
| **Indian, no (%)** | 1 (2.4) | 1 (5.6) | | |
| **Malay, no (%)** | 0 | 0 (0) | | |
| **Others, no (%)** | 0 | 2 (11.1) | | |
| **Diabetes, no (% with)** | 1 (2.4) | 1 (5.6) | | 0.54 |
| **Cataract, no (% with)** | 18 (43.9) | 12 (66.7) | | 0.11 |
| **PCIOL, no (% with)** | 2 (4.9) | 2 (11.1) | | 0.38 |
| | | **Pre-operation** | **Post-operation** | |
| **Upward displacement of optic Chiasm (UDOC), median (IQR), mm** | - | 6.4 (4.1) | 0.8 (1.6) | **<0.001#**, - |
| **Optic chiasm thickness (OpCT), median (IQR), mm** | - | 1.1 (0.7) | 2.3 (0.8) | **0.001#**, - |
| **BCVA, median (IQR), LogMAR** | 0.1 (0.2) | 0.3 (0.8) | 0.2 (0.2) | **0.005#**, **<0.001†**, 0.09‡ |
| **VFMD, median (IQR), dB** | 0.0 (2.0) | -6.4 (6.2) | -2.9 (4.1) | **0.003#**, **<0.001†**, **<0.001‡** |
| **PSD, median (IQR), %** | 1.4 (0.7) | 7.7 (9.1) | 3.3 (4.0) | **0.011#**, **<0.001†**, **<0.001‡** |
| **VFI, median (IQR), %** | 99.0 (2) | 83.0 (28.5) | 94.5 (9.3) | **0.006#**, **<0.001†**, **<0.001‡** |

Mann-Whitney U test was used to compare age, BCVA, VFMD, PSD, and VFI between controls and pituitary adenoma groups pre- (†) and post- surgery (‡). Wilcoxon Signed Rank Test was used to compare Upward displacement of optic Chiasm, Optic Chiasm Thickness, BCVA, VFMD, PSD, and VFI between patients with pituitary adenoma pre- and post- operation (#). $\chi^2$ test was used to compare all other variables between the two groups. **Abbreviations:** BCVA, best-corrected visual acuity; HVF, Humphrey visual field; IQR, interquartile range; LogMAR, logarithm of minimum angle of resolution; PCIOL, posterior chamber intra-ocular lens; PSD, pattern SD; VFI, Visual Field Index; VFMD, visual field mean deviation.

**Supplementary Table 2. Demographics and clinical characteristics of patients with pituitary adenoma with visual field recovery (n=5) and healthy controls (n=41)**

| Demographic and clinical characteristics | Controls | Patients with pituitary adenoma | | P-value |
|---|---|---|---|---|
| **N** | 41 | 5 | | |
| **Age, median (IQR), years** | 53.6 (13.7) | 56.4 (11.4) | | 0.27 |
| **Gender, male no (%)** | 19 (46.3) | 1 (20) | | 0.26 |
| **Ethnicity** | | | | |
| **Chinese, no (%)** | 40 (97.6) | 4 (80) | | 0.07 |
| **Indian, no (%)** | 1 (2.4) | 1 (20) | | |
| **Malay, no (%)** | 0 | 0 (0) | | |
| **Others, no (%)** | 0 | 0 (0) | | |
| **Diabetes, no (% with)** | 1 (2.4) | 0 (0) | | 0.72 |
| **Cataract, no (% with)** | 18 (43.9) | 5 (100) | | 0.02* |
| **PCIOL, no (% with)** | 2 (4.9) | 0 (0) | | 0.61 |
| | | **Pre-operation** | **Post-operation** | |
| **Upward displacement of optic Chiasm (UDOC), median (IQR), mm** | - | 8.7 (7.9) | 0.0 (0.8) | **0.04#**, - |
| **Optic chiasm thickness (OpCT), median (IQR), mm** | - | 0.8 (1.1) | 2.4 (1.3) | 0.08#, **-** |
| **BCVA, median (IQR), LogMAR** | 0.1 (0.2) | 0.2 (0.7) | 0.2 (0.2) | 0.27#, **0.03†**, 0.1‡ |
| **VFMD, median (IQR), dB** | 0.0 (2.0) | -3.7 (7.3) | -0.2 (1.9) | **0.04#**, **0.01†**, 1.0‡ |
| **PSD, median (IQR), %** | 1.4 (0.7) | 3.8 (9.0) | 1.5 (0.4) | 0.08#, **0.01†**, 0.9‡ |
| **VFI, median (IQR), %** | 99.0 (2) | 94.0 (25.5) | 99.0 (0.5) | 0.08#, **0.02†**, 0.9‡ |

Mann-Whitney U test was used to compare age, BCVA, VFMD, PSD, and VFI between controls and pituitary adenoma groups pre- (†) and post- surgery (‡). Wilcoxon Signed Rank Test was used to compare Upward displacement of optic Chiasm, Optic Chiasm Thickness, BCVA, VFMD, PSD, and VFI between patients with pituitary adenoma pre- and post- operation (#). $\chi 2$ test was used to compare all other variables between the two groups. **Abbreviations:** BCVA, best-corrected visual acuity; IQR, interquartile range; LogMAR, logarithm of minimum angle of resolution; PCIOL, posterior chamber intra-ocular lens; PSD, pattern SD; VFI, Visual Field Index; VFMD, visual field mean deviation.

**Supplementary Table 3. Demographics and clinical characteristics of patients with pituitary adenoma with no visual field recovery (n=13) and healthy controls (n=41)**

| Demographic and Clinical Characteristics | Controls | Patients with Pituitary Adenoma | | P-value |
|---|---|---|---|---|
| **N** | 41 | 13 | | |
| **Age, median (IQR), years** | 53.6 (13.7) | 58.4 (29.3) | | 0.6 |
| **Gender, male no (%)** | 19 (46.3) | 6 (46.2) | | 1.0 |
| **Ethnicity** | | | | |
| **Chinese, no (%)** | 40 (97.6) | 11 (84.6) | | 0.1 |
| **Indian, no (%)** | 1 (2.4) | 0 (0) | | |
| **Malay, no (%)** | 0 | 0 (0) | | |
| **Others, no (%)** | 0 | 2 (15.4) | | |
| **Diabetes, no (% with)** | 1 (2.4) | 1 (7.7) | | 0.6 |
| **Cataract, no (% with)** | 18 (43.9) | 7 (53.8) | | 0.7 |
| **PCIOL, no (% with)** | 2 (4.9) | 2 (15.4) | | 0.3 |
| | | **Pre-operation** | **Post-operation** | |
| **Upward displacement of optic Chiasm (UDOC), median (IQR), mm** | - | 6.3 (2.8) | 0.8 (2.2) | **0.001[#], -** |
| **Optic chiasm thickness (OpCT), median (IQR), mm** | - | 1.3 (0.6) | 2.2 (0.8) | **0.006[#], -** |
| **BCVA, median (IQR), LogMAR** | 0.1 (0.2) | 0.4 (0.8) | 0.2 (0.2) | **0.01[#], <0.001[†], 0.02[‡]** |
| **VFMD, median (IQR), dB** | 0.0 (2.0) | -7.4 (10.4) | -3.6 (2.8) | **0.03[#], <0.001[†], <0.001[‡]** |
| **PSD, median (IQR), %** | 1.4 (0.7) | 9.1 (9.1) | 4.6 (3.6) | 0.06[#], **<0.001[†], <0.001[‡]** |
| **VFI, median (IQR), %** | 99.0 (2) | 79.0 (42.5) | 91.0 (7.5) | **0.03[#]. <0.001[†], <0.001[‡]** |

Mann Whitney U- test was used to compare age, BCVA, VFMD, PSD, and VFI between controls and pituitary adenoma groups pre- (†) and post- surgery (‡). Wilcoxon Signed Rank Test was used to compare Upward displacement of optic Chiasm, Optic Chiasm Thickness, BCVA, VFMD, PSD, and VFI between patients with pituitary adenoma pre- and post- operation (#). $\chi 2$ test was used to compare all other variables between the two groups. **Abbreviations:** BCVA, best-corrected visual acuity; IQR, interquartile range; LogMAR, logarithm of minimum angle of resolution; PCIOL, posterior chamber intra-ocular lens; PSD, pattern SD; VFI, Visual Field Index; VFMD, visual field mean deviation.

**Supplementary Table 4. Pupillometric features in patients with pituitary adenoma pre- and post-surgery (n=27) compared to healthy controls (n=41)**

| Pupillometric feature | Controls | Patients with pituitary adenoma | | P-value |
|---|---|---|---|---|
| | | Pre-operation | Post-operation | |
| **Phasic-Blue, median (IQR), %** | 41.1 (12.6) | 31.9 (16.9) | 36.3 (10.9) | **0.01[#], <0.001[†],** 0.13[‡] |
| **Phasic-Red, median (IQR), %** | 37.6 (12.7) | 28.3 (19.7) | 33.2 (14.7) | **0.02[#], <0.001[†], 0.005[‡]** |
| **Max-Blue, median (IQR), %** | 56.0 (6.8) | 42.4 (18.8) | 51.6 (12.4) | **0.02[#], <0.001[†], 0.007‡** |
| **Max-Red, median (IQR), %** | 53.3 (9.8) | 39.4 (18.9) | 49.0 (14.4) | **<0.001[#], <0.001[†], 0.01[‡]** |
| **PIPR6s, median (IQR), %** | 13.6 (7.5) | 8.6 (7.8) | 13.9 (8.4) | **0.02[#], 0.001[†],** 0.83[‡] |
| **PIPR AUC 0-12s, median (IQR), %.s** | 202.5 (79.4) | 128.6 (106.8) | 192.3 (100.1) | **0.03[#], <0.001[†],** 0.08[‡] |
| **PIPR>1.7 slope, median (IQR), %/s** | -1.0 (0.6) | -0.7 (0.5) | -1.2 (0.9) | **0.001[#], 0.001[†],** 0.9[‡] |

Mann Whitney U- test was used to compare pupillometric features between controls and pituitary adenoma groups pre- (†) and post- surgery (‡). Wilcoxon Signed Rank Test was used to compare these features between patients with pituitary adenoma pre- and post- operation (#). **Abbreviations:** IQR, interquartile range; Phasic-Blue, phasic constriction to blue light; Phasic-Red, phasic constriction to red light; Max-Blue: maximum constriction to blue light; Max-Red: maximum constriction to red light, PIPR6s, post illuminance pupillary response 6 s after light offset; PIPR AUC 0–12, PIPR area under the curve; PIPR>1.7 slope, redilation slope 1.7s following light offset .

### Supplementary Figures

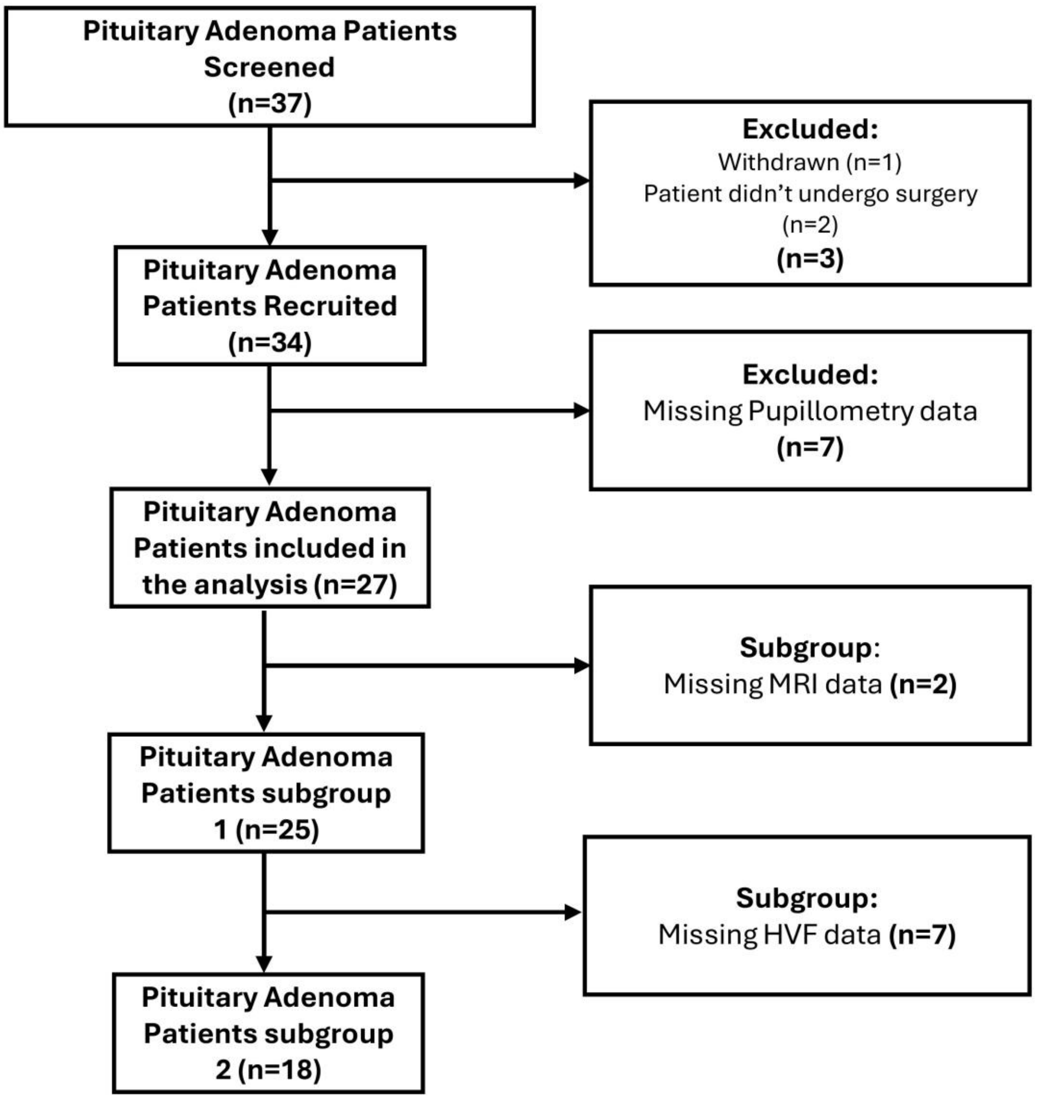


**Supplementary Figure 1. Flowchart of enrolment and exclusion of patients and data.**
**Abbreviations:** MRI, Magnetic Resonance Imaging; HVF, Humphrey Visual Field

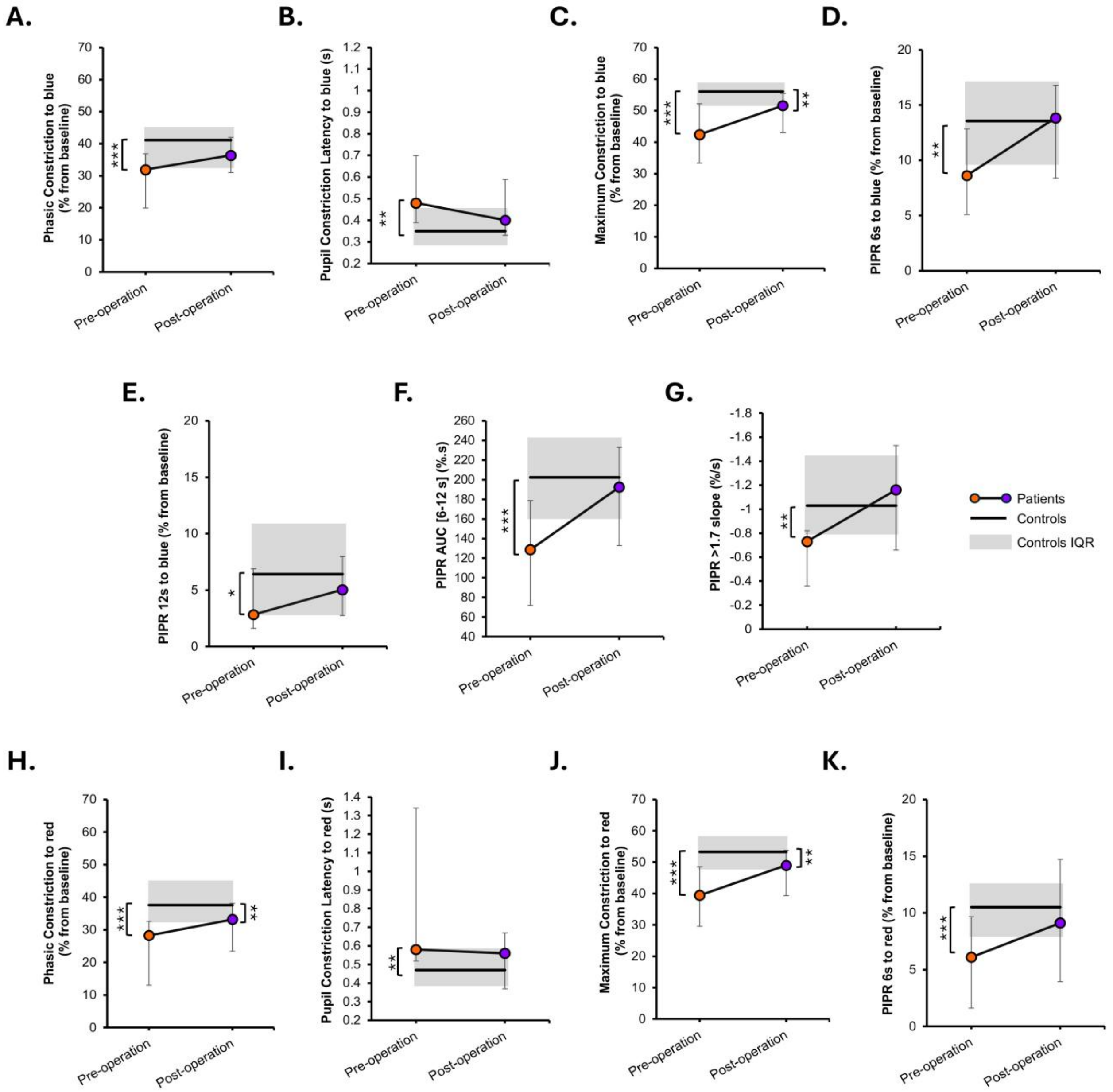


**Supplementary Figure 2. Pupillary light reflex (PLR) parameters pre- and post-surgery (n=27) compared to controls (n=41).** Each panel illustrates changes in key pupillometric features, with orange and purple markers representing individual pre- and post-operative median values, respectively. The black line represents median values of controls. Gray shaded areas indicate the IQR of control values. Error bars represent IQR within the patient group. *p < 0.05; **p < 0.01; ***p < 0.001. **Abbreviations:** IQR, interquartile range; PIPR, post illumination pupillary responses.

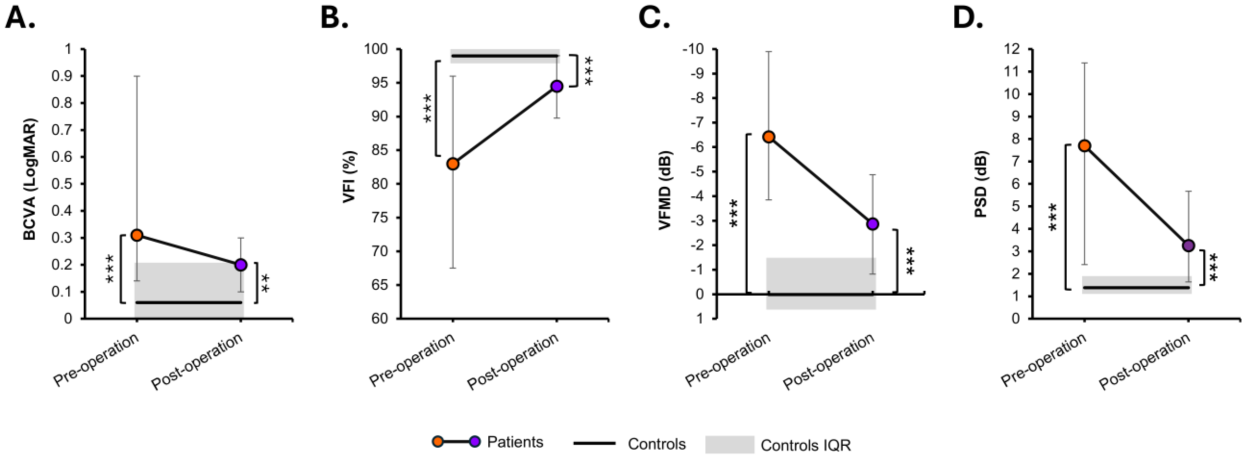


**Supplementary Figure 3. Ophthalmic assessments pre- and post- surgery (n=18) compared to controls (n=41).** Each panel illustrates changes in ophthalmic outcomes, with orange and purple markers representing individual pre- and post-operative median values, respectively. The black line represents median values of controls. Gray shaded areas indicate the IQR of control values. Error bars represent IQR within the patient group. *p < 0.05; **p < 0.01; ***p < 0.001. **Abbreviations:** IQR, interquartile range; BCVA, best-corrected visual acuity; dB, decibels; LogMAR, logarithm of minimum angle of resolution; PSD, pattern SD; VFI, visual field index; VFMD, visual field mean deviation.

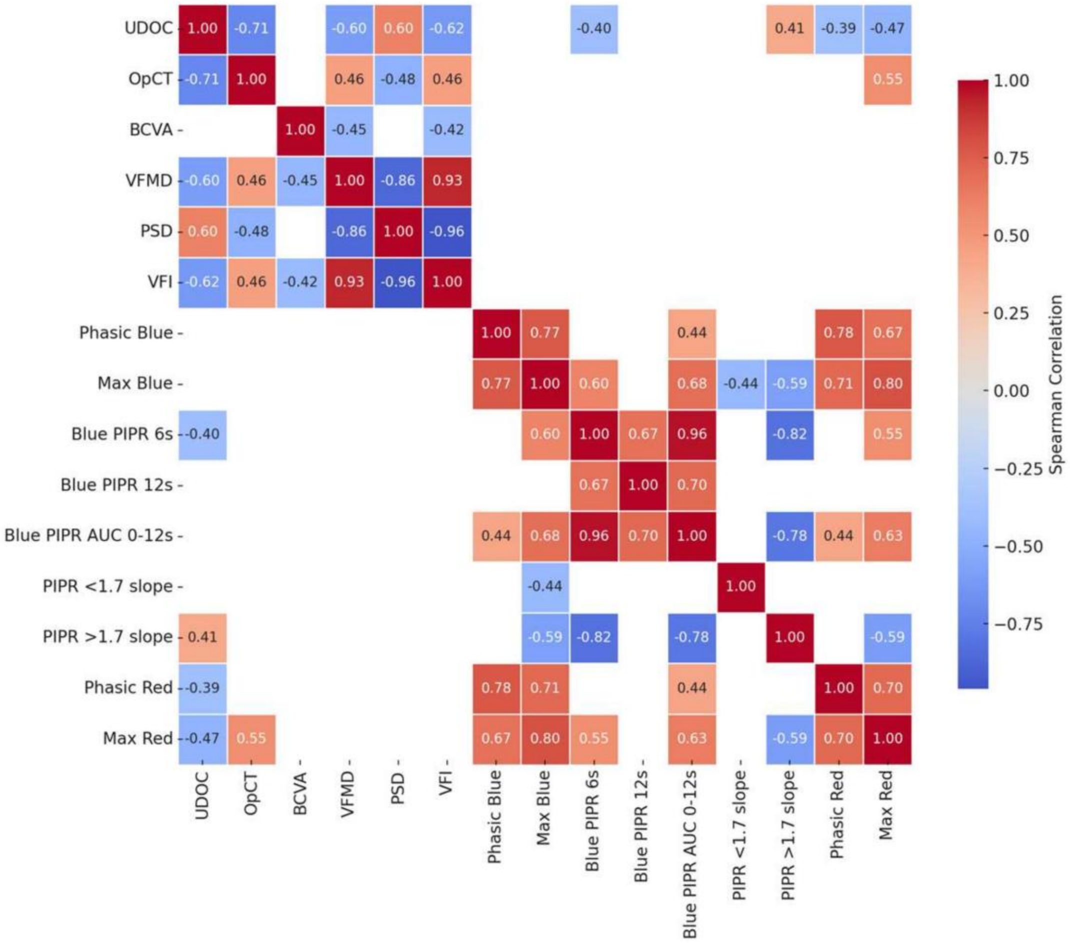


**Supplementary Figure 4. Heatmap illustrating the Spearman correlation coefficients between all measured variables** after applying the Benjamini-Hochberg (BH) correction for multiple comparisons**.** Only statistically significant correlations (*p*-adjusted < 0.05) are displayed, while non-significant cells are masked to white. Warmer colors indicate positive correlations, whereas cooler colors indicate negative correlations. **Abbreviations:** UDOC, the upward displacement of the optic chiasm; BCVA, best-corrected visual acuity; VFI, visual field index; Max Red, maximum constriction to red light; PIPR>1.7 slope, post-illumination pupillary response at >1.7 slope; OpCT, optic chiasm thickness.

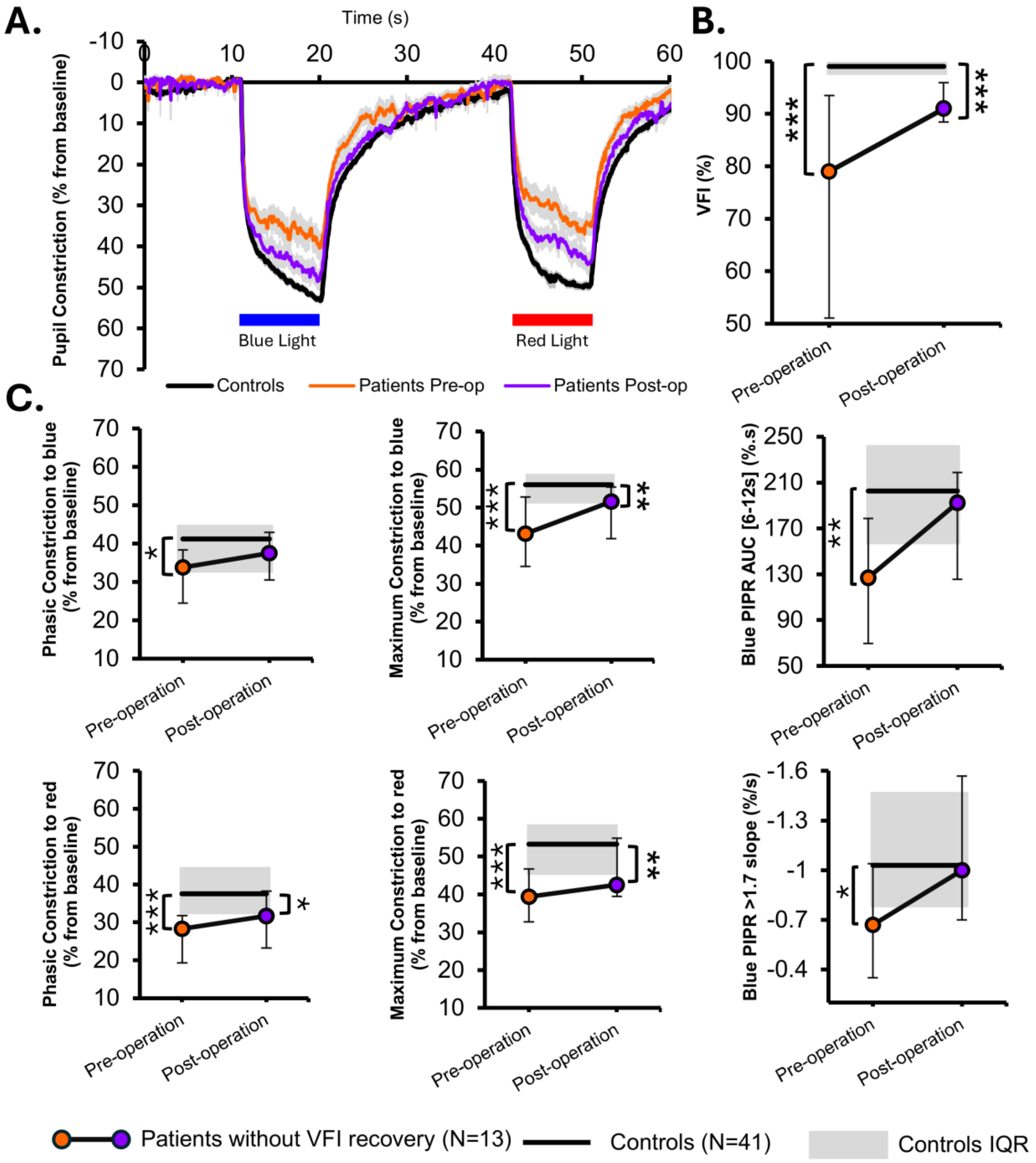


**Supplementary Figure 5. Pupillary light response and comparisons of pupillometric parameters in patients without full VFI recovery**. **A.** Mean pupil response traces to blue- and red-light stimuli, in patients with Pituitary Adenoma without VFI recovery (n=13) before (orange) and after surgery (purple) and controls (black). **B.** Difference in VFI in patients with PA before and after surgery. This sub-group of patients does not have a complete visual field recovery. **C.** Differences in main pupillometric features between patients pre- and post-operation and controls. *p<0.05; ** p<0.01; ***p<0.001. **Abbreviations:** IQR, interquartile range; VFI, visual field index; PIPR, post-illumination pupillary response; AUC, area under the curve.
